# Narcolepsy Revolution – Protocol and Methodology A diagnostic accuracy study protocol using the Dreem 3 headband for ambulatory diagnosis of narcolepsy in children and young adults

**DOI:** 10.64898/2026.03.25.26349319

**Authors:** Thomas Rossor, Carla Rush, Emily Senior, Adam Birdseye, Chiara Piantino, Laura Pérez-Carbonell, Guy Leschziner, Ullrich Bartsch, Paul Gringras

## Abstract

**Background:** Narcolepsy is a rare, lifelong neurological disorder that often begins in childhood or adolescence. Diagnosis is frequently delayed because current diagnostic testing relies on specialist in-patient sleep investigations: overnight polysomnography (PSG) followed by a multiple sleep latency test (MSLT), interpreted according to International Classification of Sleep Disorders criteria (ICSD-3-TR). These investigations are expensive, labour intensive, and available in a limited number of centres, contributing to delays and inequity of access. Automated analysis of sleep-stage probabilities (hypnodensity) using neural networks has shown promising diagnostic performance in research cohorts but still requires hospital-based PSG acquisition.

The Dreem 3 headband (DH) is a comfortable, dry-montage EEG device designed for home use. Combined with its proprietary machine learning classification of sleep stages, it may offer accurate ambulatory sleep physiology assessments and support clinical decision making.

**Methods:** This was a single-centre, prospective, observational study recruiting 60 participants aged 10 to 35 years undergoing investigation for hypersomnolence within GSTT sleep services and scheduled for PSG and MSLT as part of routine care.

Exclusion criteria included physician-diagnosed medical or psychiatric disorder that could independently account for excessive daytime sleepiness; and/ or regular use of prescribed or recreational medication known to affect sleep architecture.

Participants first wore the DH at home for five weeknights, followed by a continuous 48-hour weekend recording using two devices in rotation. They then underwent routine in-patient PSG and MSLT. PSG and MSLT were interpreted according to ICSD-3 by an experienced sleep physician and a final diagnosis determined by a sleep physiology consultant.

The primary outcome is accuracy of ambulatory DH-based assessment of sleep physiology and subsequent diagnosis of sleep disorders. We evaluate proprietary and in-house developed machine learning methods to detect SOREM epochs and predict narcolepsy diagnosis from PSG, PSG+MSLT and DH data. All algorithmic outcomes will be compared to clinical outcomes derived from current clinical standard of care.

**Discussion:** This study will provide proof-of-concept evidence for a home-based wearable EEG approach to narcolepsy diagnosis. Patient and public involvement work with young people with confirmed narcolepsy indicates high acceptability of the DH protocol: in a survey of ten young people, eight reported they would be willing to wear a sleep headband nightly at home for five days (two were unsure), and seven reported they would be willing to wear it continuously for 48 hours over a weekend (two were unsure; one said no). These findings informed the decision to restrict continuous wear to the weekend, reflecting feedback that daytime wear during school or work hours would be unacceptable. If validated, this approach could reduce delays to diagnosis, improve equity of access, and support development of a subsequent multicentre study.

**Trial registration:** IRAS Project ID: 321547. Registered October 2022. Recruitment was completed on 30 January 2026.

## Introduction

Narcolepsy is a chronic disorder of sleep–wake regulation characterised by excessive daytime sleepiness, sleep-onset rapid eye movement periods (SOREMPs), and, in type 1 narcolepsy, cataplexy. In children and young people, symptoms can be mistaken for behavioural problems, mood disorders, or ‘just being a teenager’. In a recent European study, diagnosis was delayed by a mean of 9.7 years, with younger age at symptom onset being a predictor of longer delay.^1^ Earlier identification matters because effective treatments exist but are typically only accessible once diagnosis is confirmed.

The current diagnostic pathway relies on specialist investigations: actigraphy and sleep diary, overnight PSG, and next-day MSLT. This pathway is constrained by the need for an accredited sleep laboratory and expensive hardware for accurate data collection.

In addition, specialist staff time is required for manual scoring of actigraphy, PSG and MSLT data. Manual sleep staging of PSG and MSLT is time consuming and inter-rater reliability is imperfect even under standardised criteria.^2^ The MSLT also shows biological and situational variability and can yield false-positive and false-negative results.

Machine learning based analysis of sleep neurophysiology offers a route to scalable automated diagnostics. Previous studies have shown that deep learning-based methods can reliably score sleep in diverse patient populations and identify sleep disorders, including narcolepsy with sensitivity and specificity comparable to PSG+MSLT.^3–5^

However, these approaches still depend on PSG and or MSLT data, and its acquisition in a hospital laboratory. Wearable EEG devices could remove this acquisition bottleneck by enabling prolonged home recording under ecologically valid conditions. The DH is a dry-montage wearable EEG device intended for home use, which has been validated against PSG.^6^ To our knowledge, an evaluation of the DH in a real-world clinical setting for the diagnosis of hypersomnia has not been reported.

The central scientific question for this protocol is whether narcolepsy’s characteristic sleep architecture and REM dysregulation signatures can be detected by a wearable EEG (DH) with a limited montage EEG to support accurate diagnosis at home, and whether a continuous 48-hour profile can capture daytime sleep episodes and SOREMPs in normal life settings.

### Patient and public involvement

Young people with narcolepsy and their families contributed to protocol development, including feasibility and acceptability of DH wear duration. In a structured survey of ten young people aged 14 to 19 years with confirmed narcolepsy, most respondents reported they would have preferred home sleep testing to hospital sleep laboratory testing. Eight reported they would be willing to wear a sleep headband nightly at home for five days (two were unsure), and seven reported they would be willing to wear it continuously for 48 hours over a weekend (two were unsure; one said no). Feedback also indicated that continuous wear during school or work hours would not be acceptable, which directly informed the decision to restrict continuous recording to the weekend. Assessment of feasibility was continued throughout the study and will inform interpretation and dissemination.

## Study objectives

### Primary objectives

- To determine whether ambulatory at-home monitoring using the DH can provide sufficient evidence for an accurate diagnosis of narcolepsy compared with the clinical reference standard of PSG and MSLT interpreted according to ICSD-3 criteria.

### Secondary objectives

- To determine the feasibility and acceptability of a wearable EEG device at home in a clinical context.
- Compare diagnostic accuracy using (a) up to five nights of weeknight DH data, (b) up to 48 hours continuous DH data, and (c) both combined.
- Assess whether the 48-hour continuous profile improves discrimination between narcolepsy and alternative causes of hypersomnolence, including idiopathic hypersomnia and behaviourally induced insufficient sleep syndrome.
- Explore whether ambulatory recording duration can be shortened without material loss of diagnostic accuracy.

## Methods

### Study design

This was a prospective, single-centre, observational diagnostic accuracy study.

Reporting guidance: this protocol is presented with reference to STARD 2015^7^ and STARD-AI principles; the final diagnostic accuracy report will include the completed checklist(s) as supporting information.

### Setting

Recruitment and clinical investigations took place at the Lifespan Sleep Service, Guy’s and St Thomas’ NHS Foundation Trust (GSTT), London, UK.

### Sample size

We intended to recruit 60 participants. In this clinical pathway, an approximate 50% narcolepsy diagnosis rate is expected, providing broadly balanced groups for classifier development and evaluation. The sample size was intended for proof-of-concept diagnostic accuracy and to inform design parameters for a later multicentre study.

### Eligibility criteria

Inclusion criteria: (1) age 10 to 35 years; (2) undergoing investigation for hypersomnolence within GSTT sleep services and scheduled for PSG and MSLT as part of routine care; (3) able to provide written consent or assent as appropriate.

Exclusion criteria: (1) physician-diagnosed medical or psychiatric disorder that could independently account for excessive daytime sleepiness; (2) regular use of prescribed or recreational medication known to affect sleep architecture.

### Participant identification, recruitment, and consent

Potential participants were identified sequentially from those attending sleep clinics and scheduled for PSG and MSLT. Written participant information was provided during routine care. A video consultation was offered at least two weeks later to answer questions and obtain informed consent (and parent or guardian consent where applicable) and assent for participants aged 12 to 15 years.

### Index test: Dreem 3 headband protocol

All participants received two DH devices to support the continuous weekend protocol, as recording time is limited to approximately 12 hours. Participants were trained in device use and troubleshooting. Technical support was available throughout. As this study evaluates the DH use in a ‘real world’ context, where the full protocol was not completed, the data will still be included in analysis.

Acquired headband data was uploaded to a secure server at the device manufacturer Beacon Biosignals (Boston, MA, USA) and then downloaded to secure data locations at GSTT and the University of Surrey.

Weeknight recordings: participants wore the DH overnight for five consecutive weekday nights, from bedtime until rising.

Continuous weekend recording: from bedtime on Day 5, participants wore the DH continuously for 48 hours over the weekend. Two devices were used in rotation to manage charging. Device swaps followed a pre-specified schedule to maximise continuous coverage.

### Routine clinical investigations

PSG and MSLT were recorded at the sleep laboratory facilities at GSTT using standard clinical montage and scored according to current AASM criteria^8^ by trained physiologists. MSLT followed AASM protocol with four or five nap opportunities. One or two-week actigraphy was performed prior to inpatient sleep investigations. These investigations are part of routine care.

In adult patients, PSG and MSLT data were acquired through Natus Embla NDx PSG system (Natus, USA), and analysed using Natus SleepWorks software (Natus, USA). Actigraphy data were acquired though the MotionWatch 8 actiwatch (CamNtech, UK) and analysed using MotionWare software (CamNtech, UK).

### Reference standard diagnosis

The reference standard diagnosis is a consensus clinical diagnosis made by an experienced sleep physician incorporating clinical history, actigraphy and sleep diary, overnight PSG, and MSLT interpreted according to ICSD-3 criteria.

### Outcomes

Primary outcome: diagnostic accuracy of DH-based classification for narcolepsy versus the reference standard diagnosis, reported as sensitivity, specificity, positive predictive value, negative predictive value, and area under the ROC curve.

Secondary outcomes: (1) comparative diagnostic performance for weeknight-only, continuous-only, and combined datasets; (2) acceptability and preference for home monitoring.

### Blinding

The diagnosing clinician is blinded to DH data and all machine learning outputs.

### Machine learning

We will develop custom classifiers to identify SOREM epochs and distinguish narcolepsy from other hypersomnia using EEG feature engineering and interpretable machine learning approaches.

EEG-derived features will include 1) time-domain features such as variance, Hjorth parameters activity and mobility, skew and kurtosis; 2) frequency-domain features such as absolute total power and EEG band power; and 3) nonlinear timeseries features such as permutation entropy and Fourier entropy.

Classifiers will be implemented using different approaches including Random Forest, XGBoost, and CatBoost. We will implement data splitting 80-10-10 (train-validate-test) and apply methods to balance classes across training cycles. High performing classifiers will be subjected to hyperparameter tuning to further increase performance.

The final performance of individual classifiers will be evaluated using K-fold cross-validation and AUC curves.

To further assess classifier accuracy, we will train additional classifiers on existing public domain PSG datasets that include narcolepsy patients such as the MNC dataset from the National Sleep Research Resource (sleepdata.org) and out-of-sample data Dreem headband datasets.

### Statistical analysis plan

Diagnostic accuracy will be summarised using sensitivity, specificity, and predictive values with 95% confidence intervals, and area under the ROC curve. Where appropriate, likelihood ratios will also be reported.

Agreement analyses: DH versus PSG sleep stage agreement will be assessed using epoch-by-epoch agreement metrics, including Cohen’s kappa and stage-specific agreement. Where feasible, agreement with MSLT measures will be described using correlation and classification concordance.

### Data management and confidentiality

A unique study ID was used for all DH data and analysis outputs. A separate secure linkage file connecting study ID to personal identifiers is held by the local research team. De-identified data is stored on password-protected servers for analysis in accordance with institutional policy and UK data protection requirements.

### Protocol history and amendments, ethical approval

Protocol version 1.0 23^rd^ June 2023

Protocol version 2.0 28^th^ February 2024 – Protocol modified to allow 5 nighttime recordings continuous 48-hour recordings to be undertaken in any sequence to accommodate the needs of participants.

Protocol version 3.0 17^th^ March 2025 – Inclusion criteria extended to include young people from 10 years to 35 years of age to optimise recruitment.

At time of posting, recruitment and data collection has completed (30 January 2026) and analysis is ongoing.

Ethical approval has been obtained via the London – Surrey Borders NHS Research Ethics Committee (REC, IRAS Project ID: 321547). Written informed consent has been obtained from all participants (and parent or guardian consent where participants are under 16), with written assent for participants aged 12 to 15 years.

### Dissemination

Final study results will be disseminated through peer-reviewed publication and engagement with patient organisations.

## Reporting checklist

This protocol and subsequent diagnostic accuracy manuscript will be reported in line with STARD 2015.^7^ The completed STARD 2015 checklist will be provided as Supporting information (S1 Checklist).

## Conclusion

This study evaluates whether a home-worn wearable EEG device combined with machine learning can support accurate diagnosis of narcolepsy in young people undergoing clinical investigation for hypersomnolence.

The protocol deliberately captures both weekday nocturnal sleep and an extended continuous weekend profile, aiming to increase the opportunity to observe REM dysregulation and sleep-onset patterns in normal life settings.

If DH-based classification shows strong agreement with the current reference standard, it would support the feasibility of a more scalable diagnostic pathway, potentially reducing delays, improving equity of access, and lowering the burden of repeated in-patient testing. The study is not designed to replace clinical judgement. Instead, it aims to establish whether ambulatory wearable EEG can provide clinically useful diagnostic information and to generate the parameters needed for a subsequent multicentre evaluation.

## Data Availability

This is a protocol manuscript. Once the data have been compiled, all data produced in the present study will be available upon reasonable request to the authors

## Acknowledgements

We thank the young people and families who contributed to the patient and public involvement work for their thoughtful input. We are grateful to Narcolepsy UK for consultation and support, and to the sleep physiology and clinical teams at GSTT for protocol development and study coordination.

## References

1. Zhang Z, Dauvilliers Y, Plazzi G, et al. Idling for Decades: A European Study on Risk Factors Associated with the Delay Before a Narcolepsy Diagnosis. Nat Sci Sleep. 2022;14(null):1031–1047. doi:10.2147/NSS.S359980

2. Danker-Hopfe H, Anderer P, Zeitlhofer J, et al. Interrater reliability for sleep scoring according to the Rechtschaffen & Kales and the new AASM standard. J Sleep Res. 2009;18(1):74–84. doi:10.1111/J.1365-2869.2008.00700.X

3. Stephansen JB, Olesen AN, Olsen M, et al. Neural network analysis of sleep stages enables efficient diagnosis of narcolepsy. 2018;9(1):1–15. https://www.nature.com/articles/s41467-018-07229-3. Accessed October 10, 2022.

4. Wang J, Zhao S, Zhou Y, et al. Narcolepsy Diagnosis With Sleep Stage Features Using PSG Recordings. IEEE Trans Neural Syst Rehabil Eng. 2023;31:3619–3629. doi:10.1109/TNSRE.2023.3312396

5. Vilela M, Tracey B, Volfson D, et al. Identifying time-resolved features of nocturnal sleep characteristics of narcolepsy using machine learning. J Sleep Res. 2024;33(6):e14216. doi:10.1111/jsr.14216

6. Arnal PJ, Thorey V, Debellemaniere E, et al. The Dreem Headband compared to polysomnography for electroencephalographic signal acquisition and sleep staging. Sleep. 2020;(11):1–13. https://pubmed.ncbi.nlm.nih.gov/32433768/. Accessed August 18, 2020.

7. Bossuyt PM, Reitsma JB, Bruns DE, et al. STARD 2015: an updated list of essential items for reporting diagnostic accuracy studies. BMJ. 2015;351:h5527. doi:10.1136/bmj.h5527

8. Troester MM, Quan SF, of Sleep Medicine AA, Berry RB. The AASM Manual for the Scoring of Sleep and Associated Events, Version 3. American Academy Of Sleep Medicine; 2023. https://books.google.co.uk/books?id=uKSczwEACAAJ.

